# Temperament and Psychopathology: Time-varying associations from infancy to adolescence

**DOI:** 10.1101/2025.10.15.25338125

**Authors:** Dashiell D. Sacks, Asja Abron, Pauline Vartany, Charles A. Nelson, Michelle Bosquet Enlow

## Abstract

**Background:** Temperament traits, which reflect early emerging individual differences in reactivity and regulation, are well-established correlates of psychopathology. However, studies have historically examined static temperament-psychopathology associations within limited age ranges. Research is required to understand the developmental dynamics of these associations.

**Methods:** We leveraged data from a longitudinal cohort (*N* = 767) with repeated measures in infancy and at ages 2 years, 3 years, 5 years, 7 years, and 11 years to examine predictive and concurrent associations between temperament traits (negative affectivity, surgency, effortful control, behavioral inhibition) and psychopathology (internalizing, externalizing) symptoms. We estimated time-varying associations using generalized additive mixed models to quantify variation in the significance and magnitude of associations from infancy through early adolescence.

**Results:** Greater negative affectivity consistently predicted higher internalizing and externalizing symptoms from infancy through 11 years. Surgency showed differential patterns, with higher surgency associated with lower internalizing symptoms but greater externalizing symptoms. Surgency from 2 years was associated with both internalizing and externalizing symptoms over proximal developmental intervals, whereas at 3 years, associations with externalizing extended distally through 11 years, while associations with internalizing remained proximal. Higher effortful control was associated with fewer internalizing and externalizing symptoms, with stronger effects for externalizing symptoms. Behavioral inhibition at 3 years was associated with internalizing symptoms ages 3 and 5 years. The significance and magnitude of associations between temperament domains and psychopathology symptoms varied based on developmental timing.

**Conclusions:** Temperament traits show differential associations with psychopathology symptoms, depending on the specific temperament trait and psychopathology domain. Further, the significance and magnitude of associations vary based on developmental timing. These findings highlight the importance of considering differential traits, domains, and developmental timing when considering the potential role of temperament in psychopathology.

---

Temperament traits – individual differences in reactivity and self-regulation – emerge early in life and are shaped over time by genetic, biological, and environmental factors (Putnam et al., 2024; Shiner et al., 2012). Contemporary models commonly group temperament into three core domains: negative affectivity, surgency, and effortful control (Rothbart, 2007). Research over the past several decades has positioned these traits as some of the most well supported early correlates of later psychopathology. In youth, psychopathology is often characterized by transdiagnostic symptoms that cluster along internalizing and externalizing dimensions (Conway et al., 2022; Sawrikar et al., 2022). A growing body of work has investigated associations between temperament traits and both internalizing and externalizing symptoms and disorders. However, much of the existing literature has focused on single temperament domains and narrow developmental windows, particularly in early to middle childhood. Less is known about how associations between temperament and psychopathology evolve across childhood and adolescence, and whether there are sensitive periods in which particular traits confer heightened risk for psychopathology and may most effectively inform early identification and prevention efforts.

Negative affectivity is among the most widely studied temperament traits in developmental psychopathology. The construct was first formalized by Watson and Clark (1984) and represents the tendency to experience negative emotions, such as anger, fear, frustration, and sadness (Rothbart & Bates, 2006). Negative affectivity is closely associated with internalizing (i.e., anxiety, depressive) symptoms, with numerous studies demonstrating associations both cross-sectionally and longitudinally (Behrendt et al., 2020; Flückiger et al., 2025; Lonigan et al., 2003; Scheper et al., 2017). Negative affectivity is also associated with externalizing symptoms, such as disobedience, lying, and aggression, in toddlers and young children (Eisenberg et al., 2000; Northerner et al., 2016). Twin research demonstrates that negative affectivity shares both common genetic and environmental influences with both internalizing and externalizing symptoms in childhood (Mikolajewski et al., 2013), which may help explain the common co-occurrence across symptom domains and further supports negative affectivity as a transdiagnostic marker of risk for psychopathology.

Associations between surgency and psychopathology are less straightforward. Surgency constitutes activity level, approach tendencies, impulsivity, and the expression of high-intensity positive affect, particularly in response to novel or stimulating contexts (Rothbart & Bates, 2006). Broadly, evidence suggests that higher surgency can be protective for internalizing symptoms (Delgado et al., 2018; Oldehinkel et al., 2004) but also confers increased risk for externalizing psychopathology (Dollar & Stifter, 2012; Heinze et al., 2025; Zastrow et al., 2018). Furthermore, there is some evidence that the strength and direction of these associations may vary by context and developmental stage. For example, higher levels of surgency in infancy have been linked to better later self-regulation, whereas surgency measured later in toddlerhood has been associated with poorer self-regulation in preschool (Foss et al., 2022). However, most studies examining surgency and psychopathology have focused on isolated developmental stages or used static models that do not capture how associations may change over time, and relatively little is known about how links between surgency and psychopathology vary across broader spans of development.

Effortful control encompasses the ability to voluntarily regulate attention, emotion, and behavior, including the capacity to inhibit a dominant response and to flexibly shift or sustain attention in response to contextual demands (Rothbart & Bates, 2006). The majority of evidence suggests that greater effortful control plays a protective role against the development of both internalizing and externalizing psychopathology (Delgado et al., 2018; Heinze et al., 2025; Oldehinkel et al., 2004; Scheper et al., 2017). As a regulatory domain of temperament, effortful control is often considered in the context of other temperament domains. For example, Raines et al. (2023) found that among clinically anxious children aged 8-12 years, higher effortful control protected against the effects of negative affectivity on anxiety and depressive symptoms. However, in a non-clinical community sample of children aged 3–6 years, Delgado et al. (2018) found that higher effortful control amplified the association between negative affectivity and internalizing symptoms. Effortful control begins to emerge in toddlerhood and is preceded by orienting/regulation in infancy – a developmental precursor that reflects early capacities for attentional focusing and soothability (Rothbart & Bates, 2006). However, less is known about how early orienting/regulation is directly associated with later psychopathology or when effortful control becomes a reliable predictor for later psychopathology.

Although there is substantial evidence linking early temperament traits to later psychopathology, the extant literature is limited. Most studies rely on cross-sectional data or longitudinal models that assume static associations throughout development and that focus on singular temperament traits or psychopathology domains. Childhood is a dynamic period characterized by rapid and often non-linear changes in psychological systems that may influence when and how specific temperament traits confer risk for internalizing or externalizing symptoms. These dynamic associations may be particularly important during early and middle childhood, when regulatory traits are developing and sensitive periods are thought to confer lasting developmental impact. Time-varying modeling approaches can be used to flexibly estimate how the magnitude and direction of associations between variables shift throughout development. For example, Lee et al. (2023) leveraged time-varying effects models to quantify time-varying associations between maternal childhood adversity and child psychopathology and investigate the developmental windows when intergenerational influences become salient. This approach can be used to examine whether specific temperament traits are differentially associated with internalizing and externalizing symptoms across childhood and to identify potential developmental windows during which these associations are strongest.

## The Current Study

In the current study, we used generalized additive mixed models (GAMMs) to estimate time-varying associations between temperament traits and psychopathology from infancy to early adolescence. Specifically, we quantified the associations between temperament traits and psychopathology domains as smooth functions of age, evaluating how associations change throughout childhood. We leveraged data from a large, longitudinal cohort with repeated measures of temperament collected at infancy, 2 years, 3 years, 5 years, 7 years, and 11 years of age, with psychopathology data collected from age 3 years and onward. We had two primary aims: a) to examine predictive associations between temperament traits at infancy, 2 years, and 3 years and psychopathology at later timepoints, and b) to examine time-varying associations between concurrent measures of temperament and psychopathology from ages 3 years through 11 years. This design enabled concurrent models to capture developmental change in associations and predictive models to test the predictive capacity of temperament at distinct early ages for later psychopathology.

In both aims, we focused on the three core temperament domains – negative affectivity, surgency, and effortful control (or orienting/regulation in infancy) – and on internalizing and externalizing psychopathology domains. We estimated both individual predictor models and combined models to assess the unique and shared contributions of each temperament trait over time. In addition, at age 3 years, behavioral inhibition was measured via a standardized behavioral task and was considered here as an additional independent predictor at this timepoint. Behavioral inhibition is a widely studied temperament profile, characterized by high negative affectivity and low surgency, and has been identified as an early risk factor for internalizing symptoms, particularly anxiety (Fox et al., 2023). We hypothesized broadly that negative affectivity would be associated with greater internalizing and externalizing symptoms, surgency would relate to greater externalizing and lower internalizing with potential developmental variation, and effortful control would be associated with lower psychopathology across domains. At age 3 years, higher behavioral inhibition was expected to predict internalizing symptoms only. Given the novelty of our approach, we anticipated developmental variation in associations but did not make specific hypotheses regarding their timing

## Methods

### Participants

Participants were recruited at infancy from a registry of local births in the Boston Area, MA, USA, comprising families who had indicated willingness to participate in developmental research. Families in the current analyses participated in a prospective longitudinal study to examine the development of emotion processing. Exclusion criteria included known prenatal or perinatal complications, maternal use of medications during pregnancy that may significantly impact fetal brain development (i.e., anticonvulsants, antipsychotics, opioids), pre- or post-term birth (±3 weeks from due date), developmental delay, uncorrected vision difficulties, and neurological disorder or trauma. After enrollment, families were no longer followed, and their data were excluded from analyses if the child was diagnosed with an autism spectrum disorder, or a genetic or other condition known to influence neurodevelopment. In the current analyses, participants were required to have contributed temperament and psychopathology data to be included at a timepoint. There were *N* = 689 participants at infancy (4-12 months), *N* = 513 at 2 years, *N* = 464 at 3 years, *N* = 436 at 5 years, *N* = 388 at 7 years, and *N* = 205 at 11 years, for a total analytic sample of *N* = 767 participants.

### Procedures

Parents (97% the child’s mother) were asked to complete questionnaires via an online survey prior to or during an in-person study visit. Relevant data collection for the present study included demographics, temperament, and mental health measures. The Institutional Review Board at Boston Children’s Hospital approved all study methods and procedures, and parents provided written informed consent prior to the initiation of study activities. At ages 7 and 11 years, the children also provided written assent prior to assessment.

### Measures

#### Sociodemographic Characteristics

Sociodemographic characteristics were collected during the initial visit in infancy and included child age, sex, race, ethnicity, parental education, and annual household income. Child age was also collected at each subsequent visit. Sociodemographic characteristics are reported in Table S1.

#### Child Temperament

Temperament was measured longitudinally with age appropriate questionnaires that share a common three-factor structure: negative affectivity, surgency/extraversion, and effortful control (orienting/regulation in infancy). In infancy, parents completed the 91-item Infant Behavior Questionnaire – Revised, Short Form (IBQ-R-SF; Putnam et al., 2014), rating how often their infant displayed specific behaviors during the previous two weeks on a 7-point scale (1 = never to 7 = always). Cronbach’s α values in the present sample were α = .86 for negative affectivity, α = .92 for surgency, and α = .81 for orienting/regulation. At ages 2 and 3 years, parents completed the 107-item Early Childhood Behavior Questionnaire – Short Form (ECBQ-SF; Putnam et al., 2010). Cronbach’s α values were: negative affectivity α = .82, surgency α = .84, effortful control α = .87 at 2 years; negative affectivity α = .85, surgency α = .82, effortful control α = .84, at 3 years. At ages 5 and 7 years, parents completed the 36-item Children’s Behavior Questionnaire – Very Short Form (CBQ-VSF; Putnam & Rothbart, 2006). Cronbach’s α values were: negative affectivity α = .70, surgency α = .74, effortful control α = .72, at 5 years; negative affectivity α = .74, surgency α = .75, effortful control α = .75 at 7 years. At 11 years, temperament was indexed with the 103-item Early Adolescent Temperament Questionnaire – Revised (EATQ-R; Capaldi & Rothbart, 1992) on a 5-point scale (1 = almost never to 5 = almost always). Cronbach’s α values were α = .80 for negative affectivity, α = .81 for surgency, and α = .90 for effortful control. For each scale, item scores were summed and averaged to create scale scores, with higher scores indicating greater levels of that temperament dimension. At 3 years, *N* = 370 participants completed a battery of behavioral tasks adapted from Fox et al. (2001) to measure behavioral inhibition. Children were exposed to a series of novel social and nonsocial stimuli (e.g., stranger approach, unfamiliar objects, unpredictable robot) while their latency to approach, time spent in proximity to their mother, and task completion were coded from video recordings. A composite behavioral inhibition score was calculated by averaging standardized scores across tasks, with high interrater reliability (ICC = 0.78–1.00). Full task details are reported in prior studies (Kelsey et al., 2024, Bosquet Enlow et al., *under review*) and are provided in the Supplementary Information.

#### Child Psychopathology Symptoms

##### ITSEA (3 years)

The Infant-Toddler Social and Emotional Assessment (ITSEA) was administered at age 3 years to measure child internalizing and externalizing symptoms (Carter et al., 2003). The ITSEA is a parent-report questionnaire that assesses socioemotional problems in early childhood. The ITSEA provides composite scores for internalizing symptoms based on the following scales: depression/withdrawal, general anxiety, separation distress, and inhibition to novelty. The ITSEA provides composite scores for externalizing symptoms based on the following scales: activity/impulsivity, defiance, and peer aggression. The parent rates individual items on a 3-point scale (0 indicating “Not True/Rarely,” 1 indicating “Somewhat True/Sometimes,” 2 indicating “Very True/Often”), which are then summed to calculate raw scores. Raw scores were normed by child age and gender and expressed as standard T-scores with a mean of 50 and a standard deviation of 10. Internal consistency in this sample was α = .82 for internalizing symptoms and α = .79 for externalizing symptoms.

##### CBCL (5 years, 7 years, 11 years)

The Child Behavior Checklist (CBCL) for ages 1.5 to 5 years was administered at age 5 years, and the CBCL for ages 6 to 18 years was administered at ages 7 and 11 years to measure child psychopathology symptoms. The CBCL 1.5-5 comprises 99 items, and the CBCL 6-18 comprises 113 items (Achenbach et al., 2001). The CBCL forms are well established, empirically supported questionnaires for assessing child psychopathology symptoms (Achenbach et al., 2001; Achenbach & Edelbrock, 1991; Achenbach & Rescorla, 2000, 2014). The CBCL forms produce scores on multiple syndrome and DSM-oriented scales, as well as higher-order symptom scores. Parents are asked to report on their children’s behavior during the past 6 months, with possible item scores ranging from 0 (‘Not True’) to 2 (‘Very True or Often True’). The current analyses focused on the Internalizing symptoms scale and the Externalizing symptoms scale. Internalizing symptoms comprises the following syndrome scales at 5 years: emotionally reactive, anxious/depressed, somatic complaints, and withdrawn. At 7 and 11 years, internalizing symptoms comprises anxious/depressed, somatic complaints, and withdrawn/depressed. Externalizing symptoms comprises attention problems and aggressive behavior at 5 years. At 7 and 11 years, externalizing symptoms comprises rule-breaking behavior and aggressive behavior. Subscale raw scores are calibrated and normed by child age and gender, with normed scores expressed as the standard T-score metric. For internalizing symptoms, Cronbach’s α in this sample was .82 at 5 years, .82 at 7 years, and .82 at 11 years. For externalizing symptoms, Cronbach’s α in this sample was .90 at 5 years, .89 at 7 years, and .88 at 11 years.

### Statistical Analysis

Statistical analyses were conducted in R 4.3.2. Descriptive statistics were computed for all demographic and main study variables. Prior to the main analyses, data were visually inspected for any extreme outliers or other data quality issues. Time-varying associations between temperament and psychopathology from infancy to adolescence were examined using generalized additive mixed models (GAMMs) fitted via restricted maximum likelihood (REML) in mgcv 1.9-1 (Wood, 2023). Two sets of analyses were conducted: predictive models, in which temperament measured in infancy, at 2 years, and at 3 years was examined in relation to later psychopathology; and concurrent models, in which time-varying temperament and psychopathology associations were estimated within the same assessment wave. Within each set of analyses, separate models were fit to examine associations between individual temperament traits and internalizing/externalizing psychopathology domains, and a combined model tested all traits simultaneously. Temperament scores were standardized within wave and psychopathology scores across waves prior to modeling. Individual GAMMs were fit with four components: (a) fixed main effects for the temperament trait(s), (b) a thin-plate spline capturing the overall smooth effect of age (32–140 months), (c) thin-plate interaction splines of age * temperament that capture the smooth, time-varying temperament trait-psychopathology associations across development, and (d) a child-specific random intercept to account for repeated assessments. To evaluate the time-varying associations between temperament traits and psychopathology domains, point estimates β(t) and 95% confidence intervals were extracted and plotted for each individual model. Effects are considered significant at ages when confidence intervals exclude zero. Rolling Wald χ² tests (df = 3) were used to evaluate the joint contribution of all traits in combined models; significance of the Wald test is marked by a solid gray bar above combined model plots. To support interpretation, we describe effects in the Results according to the following qualitative |β(t)| labels: small (< .15), modest (< .25), moderate (< .35), strong (< .50), very strong (≥ .50).

## Results

Sample sociodemographic characteristics are presented in Table S1. Children were predominantly non-Hispanic White, and parental education and annual household income indicated middle to high socioeconomic status.

### Predictive

#### Infancy

##### Internalizing

In individual models, there was a modest, significant association between greater negative affectivity in infancy and greater internalizing symptoms across the full age range (age 3 years: β = 0.22, 95% CI [0.14, 0.30]; age 11 years: β = 0.21, 95% CI [0.10, 0.32]). Surgency was not significantly associated with internalizing symptoms at any age. There was a small, significant association between lower orienting/regulation and greater internalizing symptoms from age ∼3.4 to ∼8.8 years (peak β = -0.13, 95% CI [-0.20, -0.06]). In the combined model, the overall time-varying association was significant across the full age range (peak χ²(3) = 48.5, *p* < .001), with negative affectivity remaining positively associated with internalizing symptoms across the full age range but surgency and orienting/regulation not significant at any age.

##### Externalizing

In individual models, there was a small, significant association between greater negative affectivity in infancy and greater externalizing symptoms from ages 3.2 to 11.4 years (age 3: β = 0.08, 95% CI [0.00, 0.16]; age 11: β = 0.11, 95% CI [0.00, 0.20]). Surgency was not significantly associated with externalizing symptoms at any age. There was a small, significant association between lower orienting/regulation and greater externalizing symptoms from ages 2.8 to 7.8 years (e.g., age 3: β = -0.09, 95% CI [-0.17, -0.00]; age 7: β = -0.08, 95% CI [-0.16, -0.01]), with effects attenuating toward zero by 11 years. In the combined model, the overall time-varying association was significant from 3.8 to 7.9 years (peak χ²(3) = 10.19, *p* = .02); only negative affectivity was significantly associated with externalizing symptoms in the combined model.

**Figure 1.**
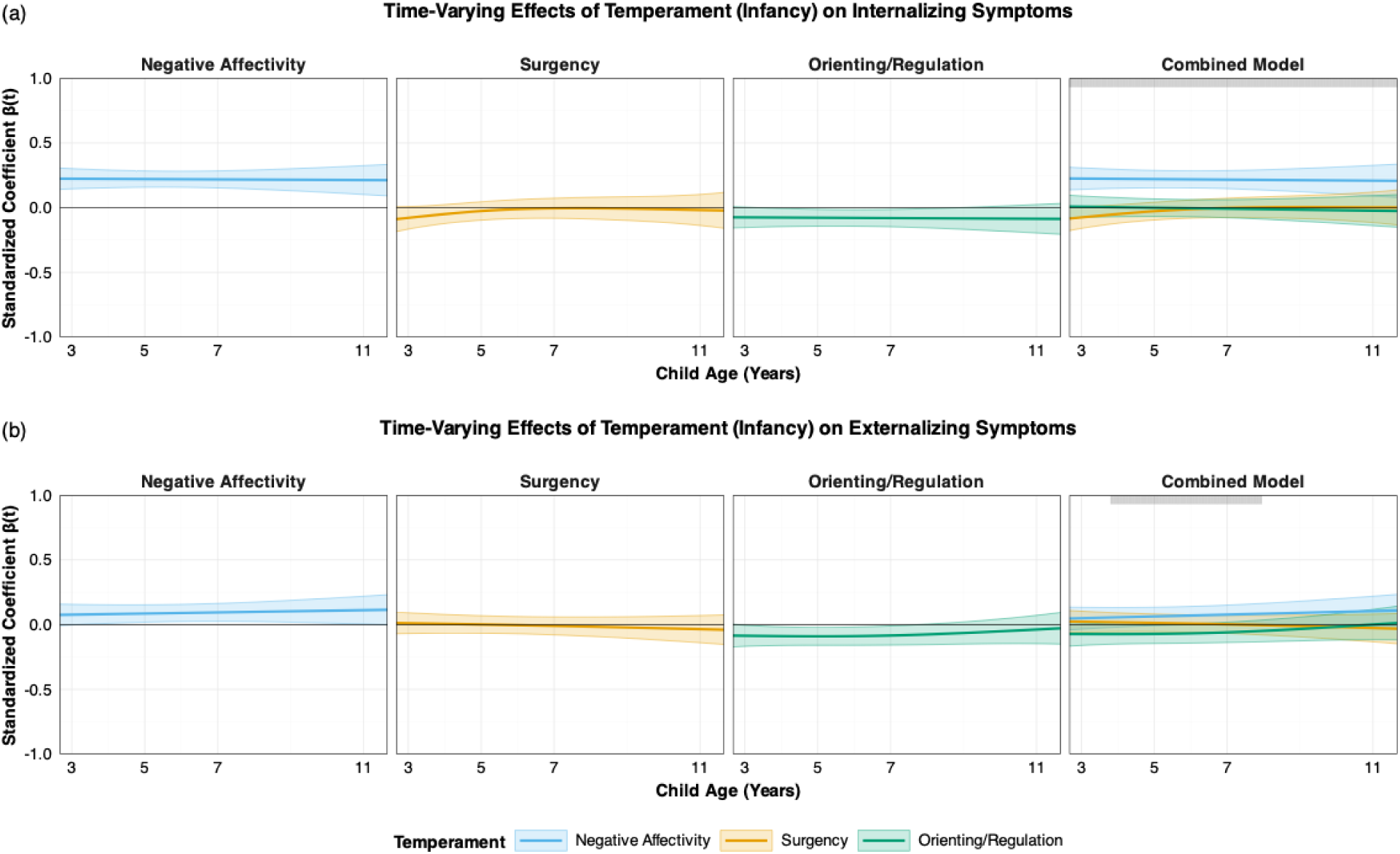
Time-varying associations between infant temperament and (a) internalizing and (b) externalizing symptoms from ages 3 to 11 years in individual (left; negative affectivity, surgency, orienting/regulation) and combined (right) models. Solid lines depict β(t) estimates for a 1-SD increase in the focal predictor, and shaded areas indicate 95% confidence intervals; associations are considered significant where the interval does not cross zero. In the combined model, significance of the set of temperament effects was tested using the Wald test, with significant ages indicated by a gray bar.

#### 2 years

##### Internalizing

In individual models, there was a strong, significant association between greater negative affectivity at age 2 years and greater internalizing symptoms starting from age 2.7, which attenuated to a more modest association by age 7, becoming non-significant by 11 years (age 3: β = 0.42, 95% CI [0.32, 0.51]; age 7: β = 0.21, 95% CI [0.13, 0.29]). There was a significant, modest association between lower surgency and greater internalizing symptoms from age 2.7 to 4.8 years (age 3: β = -0.21, 95% CI [-0.31, -0.11]) and a significant, modest association between lower effortful control at 3 years and greater internalizing symptoms from age 2.7 to 8.5 years (age 5: β = -0.22, 95% CI [-0.31, -0.13]). In the combined model, the overall time-varying association was significant from 2.7 to 8.5 years (peak χ²(3) = 100.36, *p* < .001); negative affectivity remained positively associated with internalizing symptoms from age 2.7 to 9.2 years, lower surgency was associated with greater internalizing symptoms from age 2.7 to 4.4 years, and lower effortful control was associated with greater internalizing symptoms from age 5.6 to 7.7 years.

##### Externalizing

In individual models, there was a modest, significant association between greater negative affectivity at age 2 years and greater externalizing symptoms across the full age range (age 3: β = 0.20, 95% CI [0.10, 0.29]; age 11: β = 0.17, 95% CI [0.04, 0.30]). There was a significant, small association between greater surgency and greater externalizing symptoms from age 2.7 to 7.8 years (age 3: β = 0.14, 95% CI [0.05, 0.23]; age 7: β = 0.09, 95% CI [0.02, 0.17]). There was a significant, moderate association between lower effortful control and greater externalizing symptoms starting from age 2.7 years, attenuating to a modest association by 7 years, and non-significant by 11 years (age 3: β = -0.29, 95% CI [-0.38, -0.19]; age 7: β = -0.23, 95% CI [-0.32, -0.15]). In the combined model, the overall time-varying association was significant from 2.7 to 10.7 years (peak χ²(3) = 76.11, *p* < .001); greater negative affectivity was associated with greater externalizing symptoms from age 2.7 to 7.7 years and again from 8.9 to 11.7 years, higher surgency was associated with greater externalizing symptoms from age 2.7 to 8.7 years, and lower effortful control was associated with greater externalizing symptoms from age 2.7 to 9.6 years.

**Figure 2.**
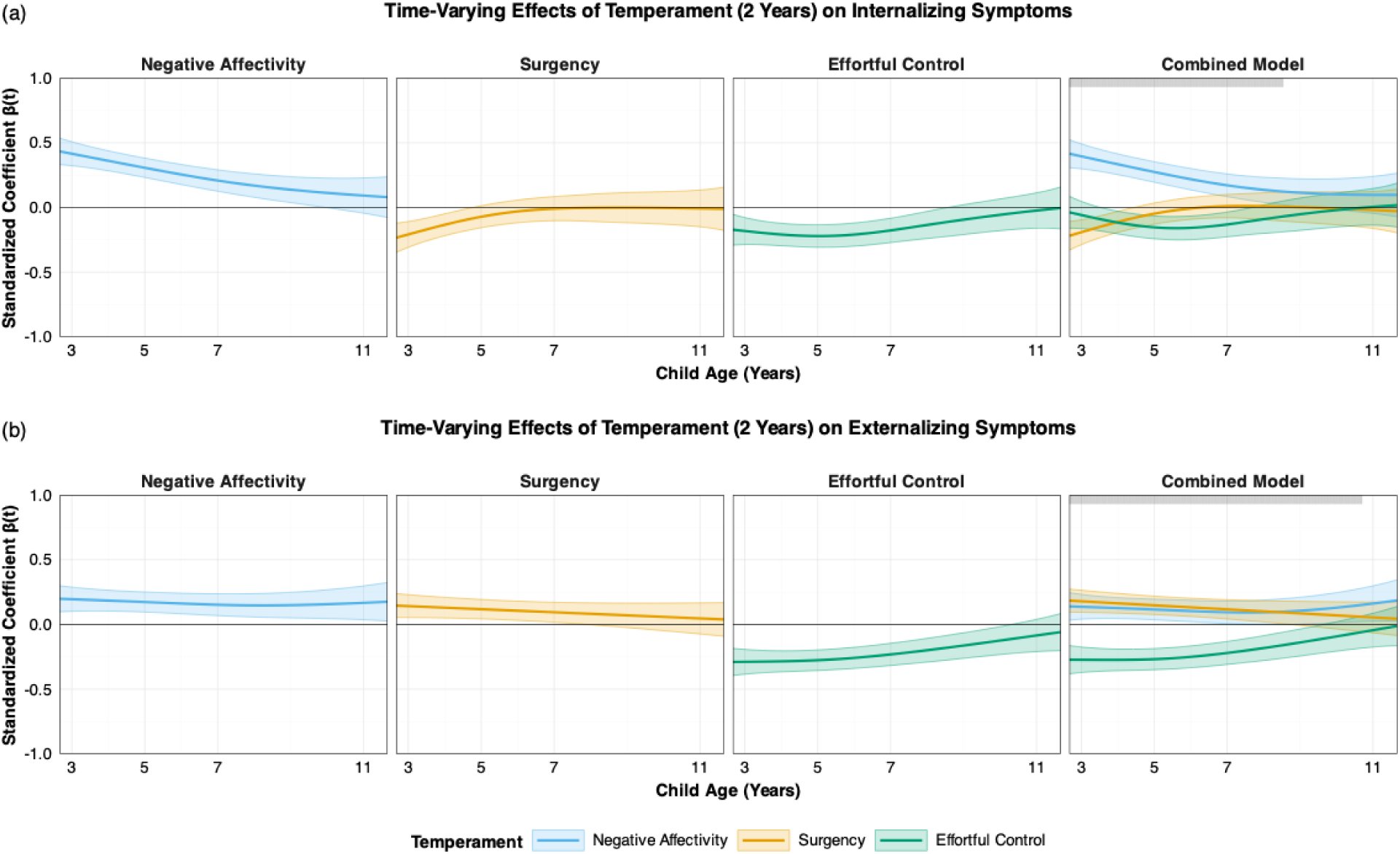
Time-varying associations between temperament at age 2 years and (a) internalizing and (b) externalizing symptoms from ages 3 to 11 years in individual (left; negative affectivity, surgency, effortful control) and combined (right) models. Solid lines depict β(t) estimates for a 1-SD increase in the focal predictor, and shaded areas indicate 95% confidence intervals; associations are considered significant where the interval does not cross zero. In the combined model, significance of the set of temperament effects was tested using the Wald test, with significant ages indicated by a gray bar

#### 3 years

##### Internalizing

In individual models, there was a significant association between greater negative affectivity at age 3 years and greater internalizing symptoms across the full age range; the association was very strong at 3 years, attenuating to a modest association by age 11 years (age 3: β = 0.55, 95% CI [0.47, 0.63]; age 11: β = 0.18, 95% CI [0.05, 0.32]). There was a moderate, significant association between lower surgency and greater internalizing symptoms from age 2.7 to 4.2 years (age 3: β = -0.31, 95% CI [-0.40, -0.21]) and a small-modest, significant association between lower effortful control and greater internalizing symptoms from age 2.7 to 8.2 years (age 5: β = -0.16, 95% CI [-0.24, -0.07]). Behavioral inhibition was positively associated with internalizing from age 2.7 to 5.8 years (age 3: β = 0.20, 95% CI [0.10, 0.30]; age 5: β = 0.12, 95% CI [0.03, 0.20]). In the combined model, the overall time-varying association was significant across the full age range (peak χ²(3) = 248.16, *p* < .001); negative affectivity remained positively associated with internalizing symptoms across the full age range, lower surgency was associated with greater internalizing symptoms from age 2.7 to 4.3 years, and effortful control was not significant.

##### Externalizing

In individual models, there was a significant association between greater negative affectivity at age 3 years and greater externalizing symptoms across the full age range; the association was moderate at 3 years, attenuating to a modest association by age 11 years (age 3: β = 0.32, 95% CI [0.24, 0.41]; age 11: β = 0.16, 95% CI [0.04, 0.27]). There was a significant association in the modest-moderate range between greater surgency and greater externalizing symptoms across the full age range (age 3: β = 0.27, 95% CI [0.18, 0.36]; age 11: β = 0.23, 95% CI [0.10, 0.36]), with effect sizes in the modest-to-moderate range. There was a strong, significant association between lower effortful control and greater externalizing symptoms from age 2.7 to 11.1 years, attenuating to a small association by age 11 (age 3: β = -0.39, 95% CI [-0.48, -0.30]; age 11: β = -0.13, 95% CI [-0.25, -0.01]). Behavioral inhibition was not significantly associated with externalizing symptoms at any age. In the combined model, the overall time-varying association was significant across the full age range (peak χ²(3) = 184.37, *p* < .001); negative affectivity, higher surgency, and lower effortful control all remained significant predictors until 7 years, with negative affectivity and effortful control attenuating to non-significance by 11 years.

**Figure 3.**
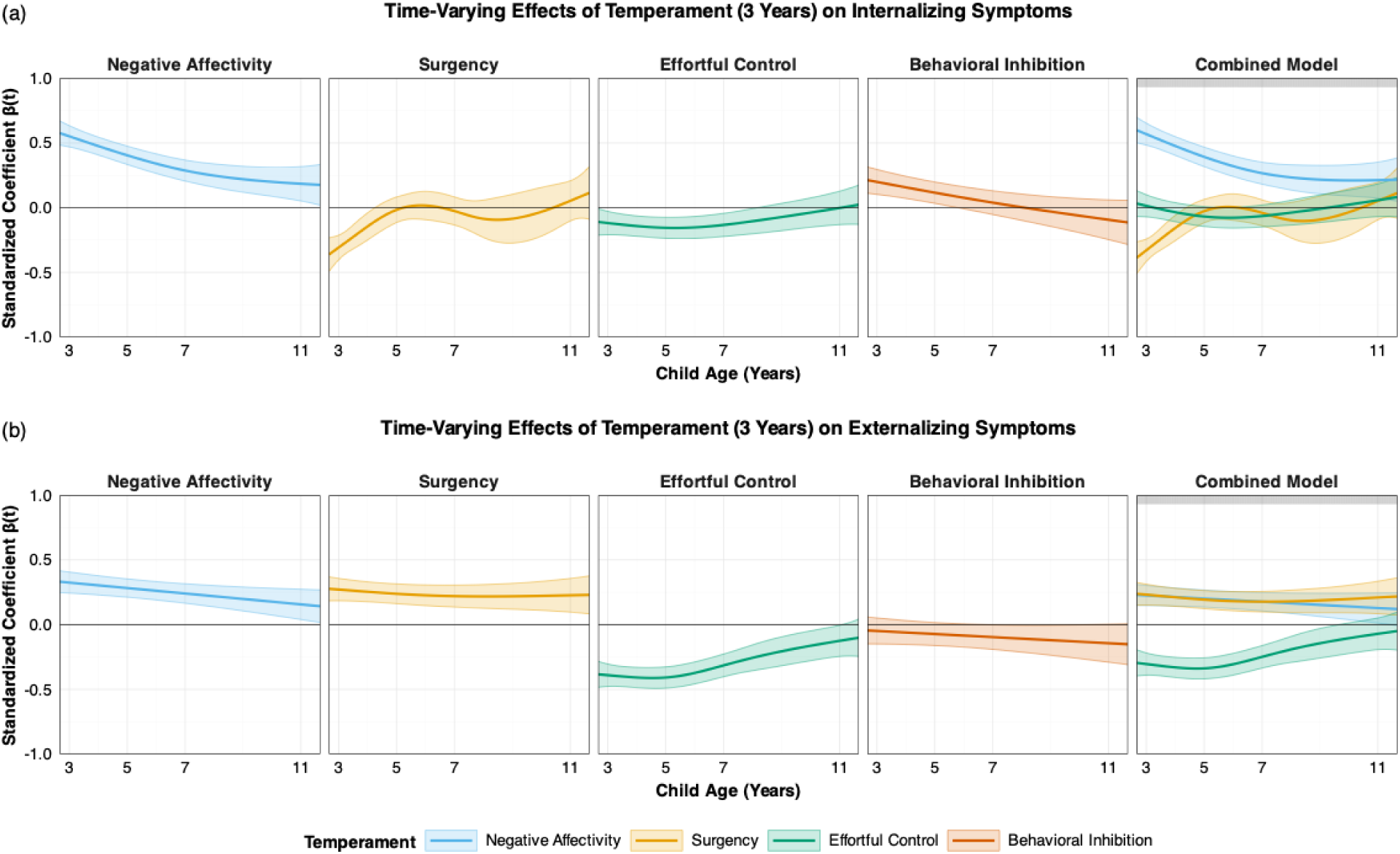
Time-varying associations between temperament at age 3 years and (a) internalizing and (b) externalizing symptoms from ages 3 to 11 years in individual (left; negative affectivity, surgency, effortful control, behavioral inhibition) and combined (right) models. Solid lines depict β(t) estimates for a 1-SD increase in the focal predictor, and shaded areas indicate 95% confidence intervals; associations are considered significant where the interval does not cross zero. In the combined model, significance of the set of temperament effects was tested using the Wald test, with significant ages indicated by a gray bar.

#### Concurrent

##### Internalizing

In individual models, there was a strong, significant association between greater negative affectivity and greater concurrent internalizing symptoms across the full age range; the association was very strong at 3 years and remained strong at 11 years (age 3: β = 0.54, 95% CI [0.48, 0.61]; age 11: β = 0.39, 95% CI [0.30, 0.48]). Lower surgency was significantly associated with greater concurrent internalizing symptoms across the full age range, with effects in the moderate-to-strong range (age 3: β = -0.33, 95% CI [-0.42, -0.25]; age 11: β = -0.35, 95% CI [-0.47, -0.23]). Lower effortful control showed a late-emerging, modest association with greater concurrent internalizing symptoms from ∼9.3 to 11.7 years (age 11: β = -0.17, 95% CI [-0.28, -0.05]). In the combined model, the overall time-varying association was significant from 2.7–11.7 years (peak χ²(3) = 444.06, *p* < .001); both greater negative affectivity and lower surgency remained associated with greater internalizing symptoms across the full age range (e.g., negative affectivity: age 3 β = 0.57, 95% CI [0.50, 0.65], age 11 β = 0.28, 95% CI [0.18, 0.38]; surgency: age 3 β = -0.34, 95% CI [-0.42, -0.26], age 11 β = -0.36, 95% CI [-0.47, -0.25]); effortful control was not significantly associated with internalizing symptoms at most ages.

##### Externalizing

In individual models, there was a strong, significant association between greater negative affectivity and greater concurrent externalizing symptoms across the full age range; the association was moderate at 3 years and strong by 11 years (age 3: β = 0.35, 95% CI [0.28, 0.41]; age 11: β = 0.45, 95% CI [0.36, 0.54]). Greater surgency was significantly associated with greater externalizing symptoms from ∼2.7 to ∼9.4 years, with effects in the modest-to-moderate range (age 3: β = 0.28, 95% CI [0.20, 0.35]; age 7: β = 0.18, 95% CI [0.11, 0.24]). Lower effortful control was significantly associated with greater externalizing symptoms across the full age range, with effects moderate and strong at 3 and 11 years (age 3: β = -0.34, 95% CI [-0.42, -0.26]; age 11: β = -0.36, 95% CI [-0.47, -0.26]). In the combined model, the overall time-varying association was significant from 2.7 to 11.7 years (peak χ²(3) = 307.63, *p* < .001); negative affectivity, higher surgency, and lower effortful control were all significant predictors at most ages (e.g., negative affectivity: age 5 β = 0.42, 95% CI [0.36, 0.49], age 11 β = 0.38, 95% CI [0.27, 0.49]; surgency: age 3 β = 0.27, 95% CI [0.21, 0.34], age 11 β = 0.08, 95% CI [-0.01, 0.17]; effortful control: age 3 β = -0.33, 95% CI [-0.40, -0.25], age 11 β = -0.31, 95% CI [-0.42, -0.20]).

**Figure 4.**
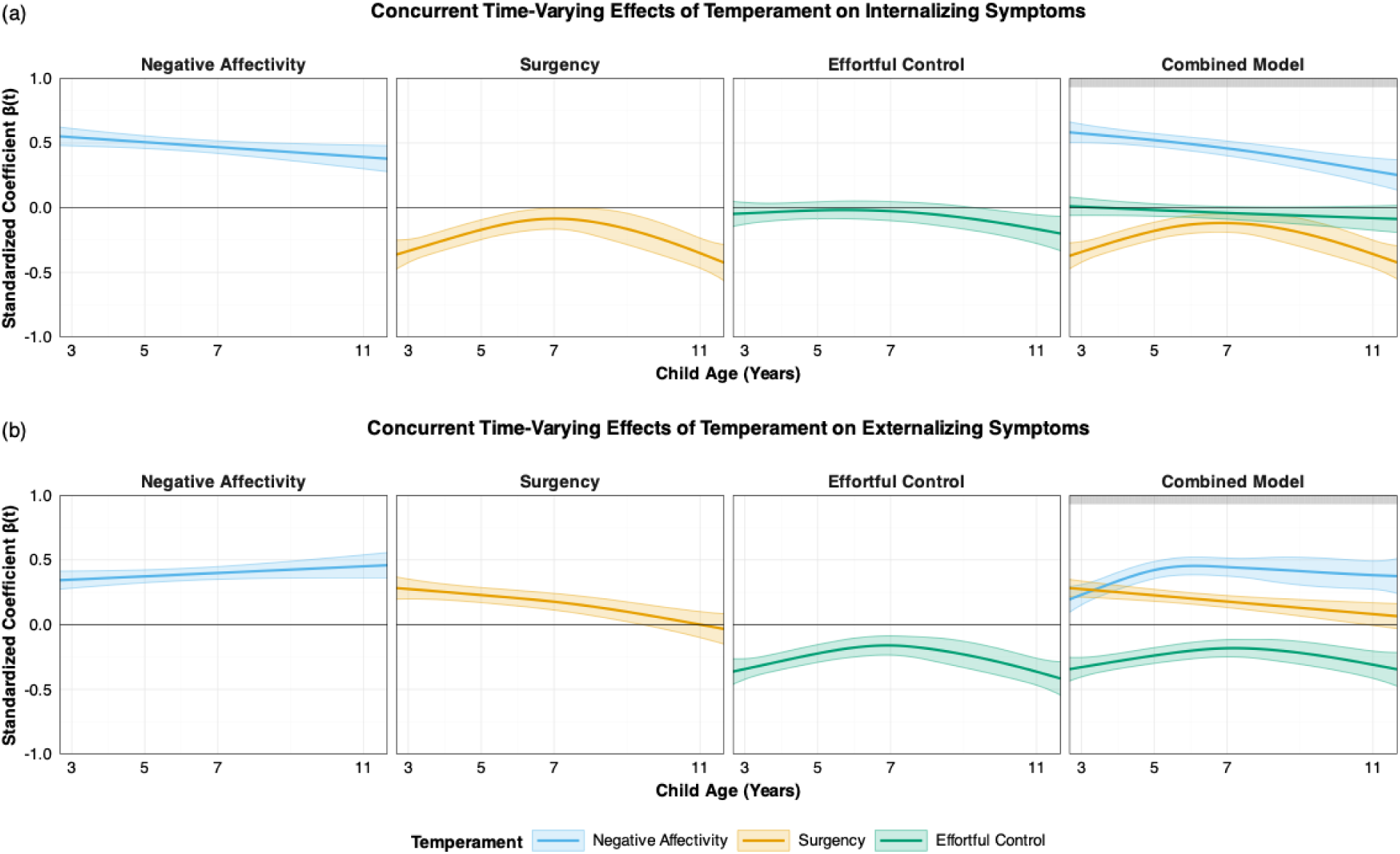
Time-varying concurrent associations between temperament and (a) internalizing and (b) externalizing symptoms from ages 3 to 11 years in individual (left; negative affectivity, surgency, effortful control, behavioral inhibition) and combined (right) models. Solid lines depict β(t) estimates, and shaded areas indicate 95% confidence intervals; associations are considered significant where the interval does not cross zero. In the combined model, significance of the set of temperament effects was tested using the Wald test, with significant ages indicated by a gray bar.

## Discussion

In the current study, we investigated associations between early temperament and child psychopathology using time-varying models in a large, prospective longitudinal cohort with repeated measures from infancy through early adolescence. We focused on the temperament traits negative affectivity, surgency, and effortful control, as well as behavioral inhibition measured at age 3, and examined their relations with internalizing and externalizing domains of psychopathology. To capture developmental dynamics, we applied a time-varying modeling approach to quantify differences in the magnitude and significance of associations across age. Leveraging our longitudinal sample, we conducted both predictive analyses, testing whether temperament measured at specific early ages (infancy, toddlerhood, preschool period) differentially predicted later internalizing and externalizing symptoms across development, and concurrent analyses, estimating how the strength of concurrent temperament–psychopathology associations changed with age. Across both predictive and concurrent models, negative affectivity emerged as the most robust predictor of psychopathology, with broad associations with both internalizing and externalizing symptoms across development. Surgency and effortful control showed more nuanced patterns, with associations differing by domain and varying in strength depending on developmental timing. Behavioral inhibition specifically predicted internalizing symptoms in early childhood only. Together, these results provide important insights into the developmental dynamics of temperament and psychopathology associations.

Negative affectivity was the temperament domain most consistently associated with psychopathology. Significant predictive associations between greater negative affectivity in early childhood and greater internalizing and externalizing symptoms through age 11 were evident from infancy. At ages 2 and 3, negative affectivity was a particularly strong predictor proximally, with associations generally remaining significant, albeit attenuated, into adolescence. These results build on decades of research that has linked higher negative affectivity to greater risk for psychopathology and provide support for negative affectivity as a transdiagnostic risk factor, with the strongest links to internalizing symptoms. Past research has demonstrated shared genetic and environmental influences between negative affectivity and both internalizing and externalizing symptoms (Mikolajewski et al., 2013). Notably, a recent study by (Morales et al., 2022) did not find an association between negative affectivity at 4 months and internalizing or externalizing symptoms measured longitudinally at ages 7, 9, or 12 years. This discrepancy may suggest that reliable associations between negative affectivity and later psychopathology emerge later in infancy (5-12 months in our study) and continue to strengthen throughout early childhood. Furthermore, the strong proximal effects in early childhood, followed by attenuated (but still significant) predictions at later ages, align with a developmental psychopathology perspective in which high negative affectivity confers early vulnerability, but diverges into multifinal outcomes as development progresses.

Surgency showed differential associations with internalizing and externalizing domains of psychopathology, with higher surgency linked to reduced internalizing symptoms but greater externalizing symptoms. Associations first emerged at 2 years, with higher surgency at ages 2 and 3 years predicting internalizing symptoms only at age 3, while only higher surgency at 3 years consistently predicted externalizing symptoms through age 11 years. These findings align with prior research suggesting that high surgency may buffer against the development of internalizing symptoms (Oldehinkel et al., 2004; Delgado et al., 2018) but increase vulnerability to externalizing symptoms (Dollar & Stifter, 2012; Zastrow et al., 2018). Our results extend prior research by demonstrating these associations are developmentally specific: Protective associations with internalizing symptoms were limited to proximal effects in toddlerhood, whereas risk associations with externalizing symptoms were persistent from preschool age onward. The lack of longitudinal associations between surgency and internalizing symptoms at the group level may reflect substantial heterogeneity in how surgency impacts risk for later internalizing symptoms. Future research may explore the contexts in which surgency confers resilience versus vulnerability. Furthermore, while our time-varying approach clarified the developmental specificity of these associations, their overall pattern remains uncertain. Future research should determine whether they reflect a continuum, with higher surgency linearly linked to lower internalizing and higher externalizing risk, or whether associations are driven by children at the extremes or by distinct subgroups showing protective versus risk patterns.

Effortful control was broadly, although modestly, protective against psychopathology, with higher levels associated with fewer internalizing and externalizing symptoms. Small associations were observed between infant orienting/regulation and later psychopathology symptoms but did not remain significant in combined models after accounting for all three temperament domains. More stable associations emerged from age 2, with lower effortful control predicting greater internalizing and externalizing symptoms, and the strongest effects evident for externalizing symptoms at age 3. These findings are consistent with prior research that has reported that higher effortful control mitigates risk for psychopathology (Oldehinkel et al., 2004; Scheper et al., 2017; Delgado et al., 2018; Heinze et al., 2025). Our results suggest that associations with externalizing symptoms are more robust. The pattern of strengthening associations of effortful control with psychopathology symptoms between infancy and age 3 is consistent with developmental models that emphasize the gradual emergence of effortful control across early childhood. Importantly, as a regulatory domain of temperament, effortful control is often considered in relation to other domains. While our combined models accounted for shared variance across traits, given the complexity and novelty of our existing models, we did not examine interactive effects directly. Future research should investigate how such interactions evolve across development and whether effortful control moderates risk conferred by other temperament domains into adolescence.

Behavioral inhibition is widely recognized as one of the most robust temperamental profiles linked to internalizing symptoms, particularly anxiety (Aktar & Pérez-Edgar, 2024; Fox et al., 2023). Therefore, we included a standardized observational measure of behavioral inhibition available at age 3 as an additional predictor in our analyses. We found that behavioral inhibition was associated only with internalizing symptoms at ages 3 and 5, with no associations observed for externalizing symptoms at any age or for internalizing symptoms later in childhood. The specificity of these findings suggests that behavioral inhibition is most relevant proximally for internalizing symptoms, while the absence of longer-term effects may reflect its modest stability across development and/or susceptibility to modification by caregiving and broader environmental contexts (Chronis-Tuscano et al., 2009; Lawrence et al., 2020; Lorenzo et al., 2022; Tang et al., 2020; Zeytinoglu et al., 2025).

The pattern of associations in concurrent models largely paralleled the predictive findings. Negative affectivity was strongly associated with both internalizing and externalizing symptoms. Surgency consistently buffered against internalizing symptoms while conferring risk for externalizing symptoms, and effortful control most reliably predicted lower externalizing symptoms. For surgency and effortful control, the magnitude of effects was stronger in early childhood and early adolescence, with weaker associations in middle childhood (5 and 7 years). These fluctuations could reflect developmental variation in the salience of temperament– psychopathology associations, or, alternatively, may be driven by methodological factors, such as differences in items and factor structure across the different temperament and psychopathology scales. Of note, we administered the very short form of the temperament measure at 5 and 7 years; thus, the attenuated associations at these ages could reflect limitations of the very short form scales.

### Strengths and limitations

The large, prospective longitudinal sample with repeated assessments from infancy through early adolescence enabled both predictive and concurrent models and lends to the robustness of our findings. At the same time, the results should be considered in the context of its limitations. Although the longitudinal design allowed us to examine developmental changes, the spacing of assessments required models to interpolate across gaps (particularly between 7 and 11 years), which may obscure more nuanced timing effects. Differences in sample size across time points influenced the width of confidence intervals (significance) at different ages and between models. The sample was predominantly White and of higher socioeconomic status, from a single region in North America, which may limit generalizability. All measures of temperament and psychopathology, aside from behavioral inhibition, relied on parent report. Finally, although we used developmentally appropriate measures of temperament and psychopathology, the forms differed across ages, making it difficult to disentangle whether observed variations reflect developmental change, differences in measurement, or a combination of both.

### Conclusions

In this study, we used a novel time-varying modeling framework to elucidate the magnitude and significance of predictive and concurrent associations between temperament (negative affectivity, surgency, effortful control, behavioral inhibition) and internalizing and externalizing psychopathology symptoms from infancy through adolescence. We found that negative affectivity was a consistent transdiagnostic risk factor from infancy. From 2 years, surgency and effortful control exhibited developmentally specific and domain-differentiated associations. Behavioral inhibition, measured only at age 3 years, specifically predicted internalizing symptoms proximally at 3 and 5 years. Overall, these findings support temperament domains as significant predictors of psychopathology from early childhood and highlight the importance of considering developmental dynamics and distinct temperament and psychopathology domains in future research.

## Data Availability

All data produced in the present study are available upon reasonable request to the authors

